# Understanding how future-fit the English pre-registration nursing, midwifery and allied health profession curricula are: A mixed methods survey

**DOI:** 10.64898/2026.11.24.26368837

**Authors:** J. Downey, M. Varga, P. Rae, M. Koroi, A. Jones, E. Golder

## Abstract

**Background:** Health education must respond to changing population needs to ensure healthcare is fit for purpose. Ageing, overwhelmed services, multimorbidity and persistent health inequalities are increasing the need for prevention and community services, whilst digital health is rapidly reshaping how care is delivered. These transformation needs are international, but in 2025 the English NHS 10 Year Plan outlined clear targets with implications for healthcare education. Despite this, little is known about how prepared the current curricula are, therefore, the aim was to explore what curricula are being delivered in English pre-registration programmes.

**Methods:** An online survey of nursing, midwifery and allied health educators in England was undertaken which included fixed-response and open-text questions related to the priority given to digital, community and preventative health compared to other elements of curricula, the volume and format of future-facing curricula, and how to modernise programmes. Data was analysed using descriptive and inferential statistics and content analysis to identify complementary findings and tensions.

**Results:** Of 166 eligible respondents, 31.3% described prevention as a strong component of their course, compared with 21.7% for community and 3.6% for digital health. Few programmes offer dedicated space to cover these elements and over 90% do not use established frameworks to guide the development of education. Open-text accounts described pressures on curriculum space and staff time, alongside the need for educator development, relevant technologies and practice-learning opportunities. Examples of established provision and helpful partnerships showed that experiences varied contingent on a complex interplay of factors.

**Conclusions:** Despite calls to align education with changing health systems, provision remains uneven. Our findings suggest that curricular change cannot be achieved by higher education institutions alone and developing future-proof education requires concurrent alignment of curriculum priorities, educator expertise, practice-learning capacity, and resources across universities, placement providers, regulators, and policymakers.

**Practice points:**

- Current education provisions indicate that most programmes are immature against the English priorities for healthcare reform.
- There is a need for a whole systems commitment to modernisation if those educating the next generation of the workforce can shift towards a “future fit” version of their professions.
- Universities must recruit an educator workforce that has the right expertise and experience to equip students with the capabilities to mobilise reform and support their current workforce with specialist support, resources, training and time.
- Universities, service providers and professional regulators have complementary roles in enabling change as each stakeholder is currently creating roadblocks for educator champions who are working in a constrained climate, stifling modernisation.

## Introduction

Health practitioner education must respond to changes in population needs and changing healthcare models. By 2030, one in six people worldwide is projected to be aged 60 years or older, with the number in this age group expected to reach 2.1 billion by 2050 [1]. Ageing populations, healthcare operational malalignment, multimorbidity and persistent health inequalities are increasing the need for continuity, prevention and coordination across health and care services. These challenges are experienced internationally [2,3] and the World Health Organisation’s (WHOs) framework for integrated people-centred health services calls for care to be organised around people throughout the life course and across settings [4]. Preparing professionals for these services is consequently an educational, as well as a service-delivery, challenge.

In England, the 2024 independent investigation of the NHS described substantial difficulties in healthcare access and performance, alongside an imbalance between hospital and community provision [5]. The 2025 NHS 10 Year Plan set out three shifts needed in healthcare provision: from hospital to community, sickness to prevention and analogue to digital and highlighted their complementary interactions [6,7]. Calls to align education with health systems are not new and fragmented education, hospital-centric, and insufficient attention to teamwork have been noted as hindering appropriate care [8]. WHO guidance has subsequently called for closer alignment between education and health services, regular curriculum renewal, support for educators and training relevant to population and service needs [9]. In 2022, WHO’s framework continued to identify alignment between health-professions education, population needs and health-system demands as a priority [10].

Yet, translating these ambitions into educational practice is not straightforward. Curriculum renewal is a substantial undertaking and may be driven by changing community and patient needs as well as regulatory changes and knowledge or skills gaps [11]. In the United Kingdom, educators continue to identify a need for placements, resources, staff development, curriculum space, collaboration and institutional support for embedding innovations effectively [12]. The maturity of contemporary models of healthcare will also differ across professions complicating attempts of widespread education change [13,12]. Digital health also brings unique considerations for workforce education given the rapid changes in professional responsibilities and disparate terminology [14,15,16]. These findings suggest that curricular alignment cannot be achieved through content or guidance alone and attention must also be given to the conditions in which educators develop and deliver curricula.

Understanding what educators are currently able to do, what they perceive is needed, and what enables or constrains further change is therefore important to translating health-system ambitions into educational practice. Little is known about the current preparedness of pre-registration education which could undermine healthcare transformation given students are the leaders of the future. The current research, therefore, aimed to examine the provision relevant to the three shifts within pre-registration education in England. The research also asked about challenges encountered or anticipated, and the support needed to develop and/or sustain provision.

## Methods

### Ethics statement

The study was approved by the University of Plymouth Faculty of Health Research Ethics and Integrity Committee (Project ID: 6650). All data collection was voluntary and signed informed consent for respondent participation and the publication of anonymised findings was collected prior to taking part via the online survey. Data was handled in accordance with the legislative and university regulations.

### Design

A mixed method, cross sectional online survey was undertaken using a previous co-designed survey as a template [12]. A pragmatic stance was adopted where quantitative and qualitative data were analysed separately as they were answering distinct elements of the research question.

### Participant recruitment

The survey was administered using Jisc Online Surveys between 3 October 2025 and 31 May 2026. Eligible participants were those who had responsibility for designing or delivering pre-registration programmes in English nursing, midwifery and allied health professions (AHPs). Recruitment used professional and higher education networks, NHS contacts, a study expert reference group, the Council of Deans of Health and professional bodies as advertisement stakeholders. Participation was voluntary and recruitment was non-probability based.

### Survey tool

The questionnaire drew on a Council of Deans of Health’s public health survey [12] and was adapted to examine curriculum provision relevant to the three shifts by sharing it with an expert advisory group for feedback. It comprised three sections. First, respondents provided descriptive information. Second, fixed-response questions examined curriculum provision across each shift, including its priority within the curriculum, use and awareness of relevant frameworks and resources, teaching formats, and the extent to which different content was included. Third, for each shift, respondents answered three open-text questions about a) the pertinent content included in the curriculum, b) challenges that may or do prevent the provision of three shift content, and c) support that has or would help improve provision of content. A fixed-response question presenting a list of potential three shift content areas followed the open text response. This provided an opportunity to capture both unprompted (free-recall) and prompted (cued-recall) descriptions of curriculum content.

### Data analysis

Quantitative findings are reported as counts and percentages, using the number of respondents answering each question as the denominator where available. Where respondents could select several answers, the categories are not mutually exclusive. Chi-Squared test and original regressions were used where appropriate to examine differences across professions. Data were analysed descriptively using MS Excel and SPSS 29 (IBM) software. Qualitative data was coded in NVivo 14 and organised into categories describing education content, challenges and support. Codes for challenges and support were developed inductively consistent with content analysis [19]. The method of analysis was agreed before analysis and coding was undertaken primarily independently, with periodic discussions. The discussions included consideration of the reasoning behind coding decisions, with codes and their descriptions refined where necessary to develop a shared understanding of meanings. Definitions and descriptions for each code were also recorded in NVivo to support consistency throughout the analysis. Quantitative results and coding categories were only brought together for a domain-based joint display to explain what fixed responses could not capture or to answer different aspects of the research question [17,20,21].

## Results

### Respondents

The analysis included 166 eligible educators. Nursing and midwifery accounted for 71 respondents (42.8%), AHPs for 94 (56.6%) and multiple programmes for one (0.6%). AHPs were categorised into small, medium and large groups based on the workforce proportions. Respondents worked across all nine English regions with a quarter based in the Southwest. Mean experience as an educator was 10.2 years (SD 7.5; range 0.5–40). Table 1 presents the demographic and professional profile of the cohort. Art therapy was the only allied health not represented in the data.

**Table 1.**
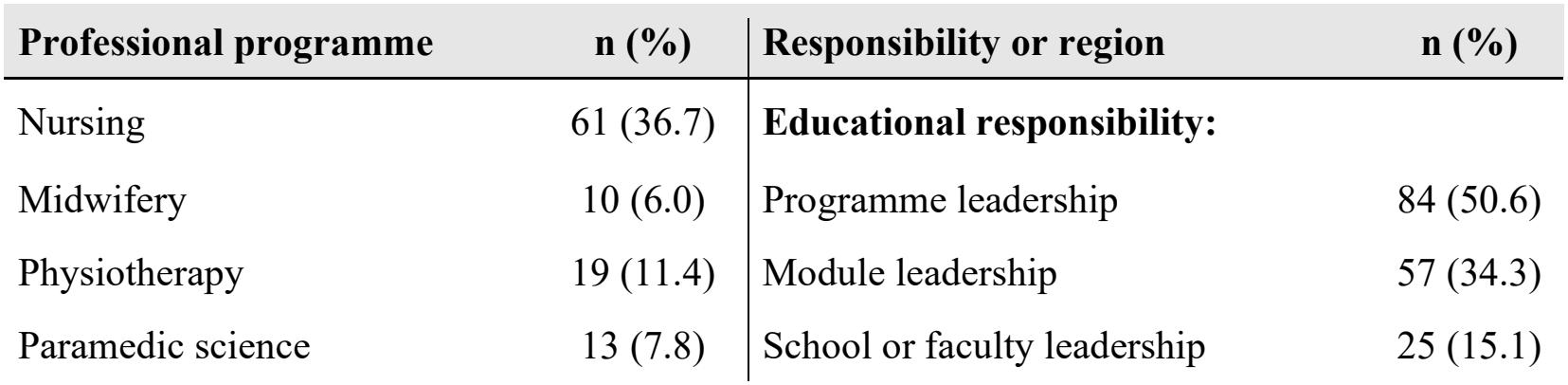
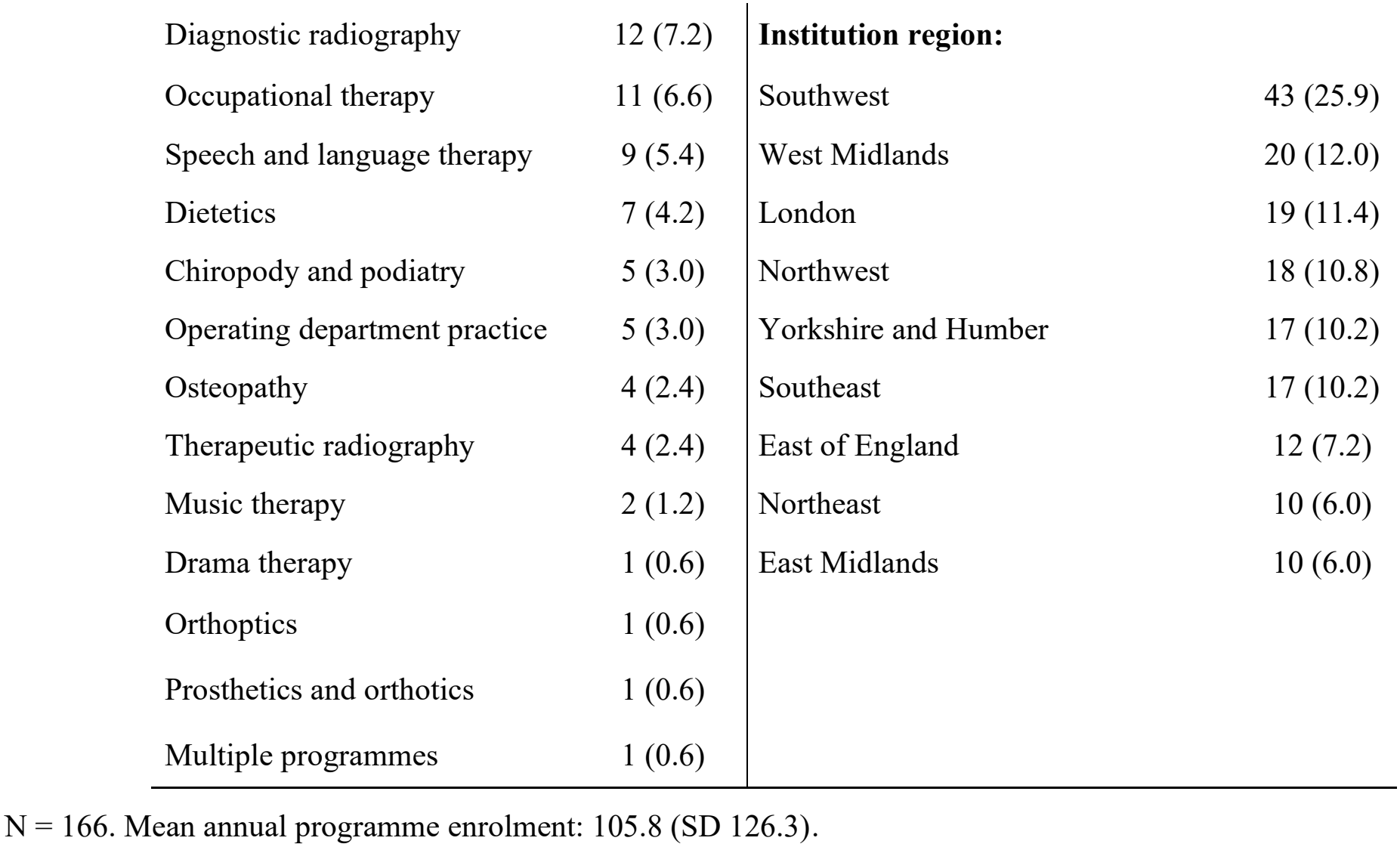
Respondent characteristics.

| Professional programme | n (%) | Responsibility or region | n (%) |
|---|---|---|---|
| Nursing | 61 (36.7) | **Educational responsibility:** |  |
| Midwifery | 10 (6.0) | Programme leadership | 84 (50.6) |
| Physiotherapy | 19 (11.4) | Module leadership | 57 (34.3) |
| Paramedic science | 13 (7.8) | School or faculty leadership | 25 (15.1) |
| Diagnostic radiography | 12 (7.2) | **Institution region:** |  |
| Occupational therapy | 11 (6.6) | Southwest | 43 (25.9) |
| Speech and language therapy | 9 (5.4) | West Midlands | 20 (12.0) |
| Dietetics | 7 (4.2) | London | 19 (11.4) |
| Chiropody and podiatry | 5 (3.0) | Northwest | 18 (10.8) |
| Operating department practice | 5 (3.0) | Yorkshire and Humber | 17 (10.2) |
| Osteopathy | 4 (2.4) | Southeast | 17 (10.2) |
| Therapeutic radiography | 4 (2.4) | East of England | 12 (7.2) |
| Music therapy | 2 (1.2) | Northeast | 10 (6.0) |
| Drama therapy | 1 (0.6) | East Midlands | 10 (6.0) |
| Orthoptics | 1 (0.6) |  |  |
| Prosthetics and orthotics | 1 (0.6) |  |  |
| Multiple programmes | 1 (0.6) |  |  |

N = 166. Mean annual programme enrolment: 105.8 (SD 126.3).

### How much of the three shifts are included and what educators need

There was a statistically significant difference between the three shifts reported in programmes (ꭕ^2^(6, N = 498) = 71.17, *p* < .001, Cramer’s V = 0.27). Prevention was most frequently rated as a strong curricular component. A small minority did not consider the shifts pertinent areas of education relative to other course content. Figure 1 shows the distribution of responses as a percentage of total number of responses per shift area. Large (OR=0.51, 95% CI [0.57-2.3], p< 0.05) and small (OR=0.26, 95% CI [0.24-1.57], p< 0.05) health professions had lower odds of reporting community as a priority, whereas nursing and midwifery had a higher odds of reporting prevention compared to large AHP professions (OR=0.37, 95% CI [0.12-0.74], p<0.05).

**Figure 1.**
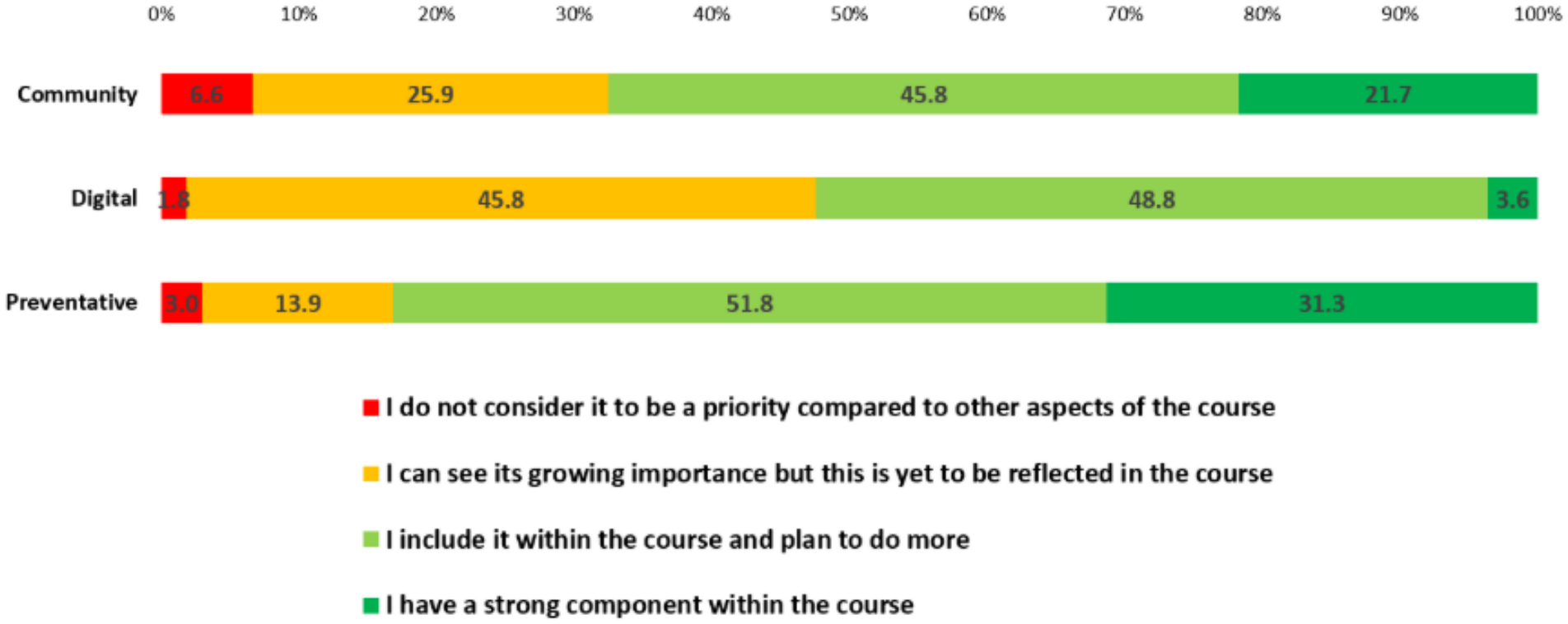
Percentage of respondents indicating the level of priority given to each shift within pre-registration programmes.

Figure 2 presents a visual representation of the shared challenges to the modernisation of education. Collectively, workforce upskilling, staff time and curriculum space were highlighted as the main challenges across all three shifts.

**Figure 2.**
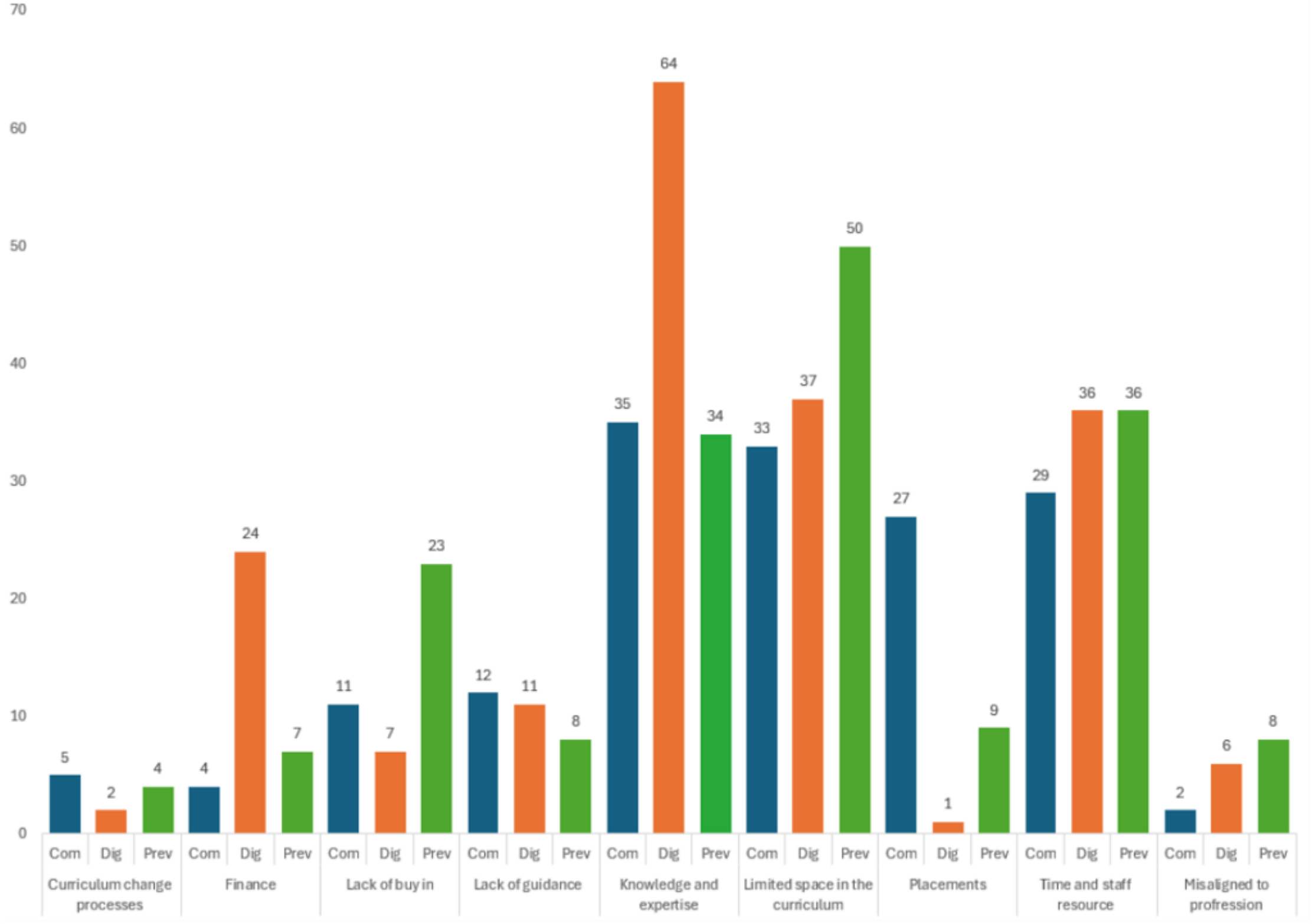
The shared challenges to the modernisation of education across the three shift areas, numbers displayed shown count of responses coded within each category.

Keeping up with advances in the topic and access to equipment was seen as a pressing challenge for digital compared to community and preventive health. However, as one respondent notes, specialist access could bolster the educator workforce.

> *“Improving collaboration with digital health experts in the university has helped inform our curricula and increase awareness of developments in healthcare*.*” (R166)*

A lack of collaboration between education and placement settings was seen as a specific challenge for community aspects of pre-registration. The data described how opportunities in services could limit what educators could offer students. One response reported an imbalance between acute and community experiences:

> *“The majority of placements learners experience are within the acute setting, more support with capacity in varied community settings would be beneficial*.*” (R144)*

Some responses also noted no specific barriers for more preventative health content but raised a question about the depth of learning possible within existing provision, even where topics were included:

> *“Only being able to introduce concepts and ideas e*.*g. behaviour change approaches, rather than train students to be skilled in them due to time constraints and a full curriculum*.*” (R62)*

Other respondents described prevention as established within teaching and practice, providing some context for the numerical contrast between prevention and digital health:

> *“Preventative health is already discussed throughout the programme as part of normal osteopathic teaching and clinical practice*.*” (R157)*

Lastly, respondents raised the issue of difficulties with changing modules and programme requirements, highlighting how the lack of agility in university processes impedes modernisation in pre-registration:

> *“Sometimes module learning outcomes are very specific and inflexible and make it difficult to adapt. Some of the things I would like to include are difficult due to the other things I am required to deliver in the module*.*” (R57)*

### How are the “three shifts” taught in programmes?

Few respondents reported using a specific framework for teaching: 15/166 (9.0%) for community, 33 (19.9%) for digital health and 30 (18.1%) for prevention. Overall, 105 (63.3%) reported no framework utilisation across the three shifts.

Open-text responses highlighted that proactive holistic care, out-of-hospital care, neighbourhood health, digital healthcare and monitoring, health information technology, artificial intelligence, public health models, health improvement and wider determinants of health are the dominant content areas covered across the three shift areas (Table 2).

**Table 2.** Summary of taught topics. Cases show the number of responses recorded under each code.

| **Domains** | **Cases** |
|---|---|
| **Digital** |  |
| AI | 51 |
| Acquiring information/knowledge management | 9 |
| Data analytics | 6 |
| Data governance | 28 |
| Digital creation and innovation | 6 |
| Health inequalities and ethics | 21 |
| Digital healthcare and monitoring | 99 |
| Digital identity | 14 |
| Digital key professional duties | 1 |
| Digital literacy and communication | 45 |
| Digital pedagogy | 34 |
| Digital placements | 1 |
| Digital information and technology | 65 |
| **Community** |  |
| Wider determinants of health, inequalities and community assets | 21 |
| Risk management | 12 |
| Proactive holistic care | 49 |
| Out of hospital care | 42 |
| Neighbourhood health | 31 |
| Local healthcare management | 13 |
| **Preventative** |  |
| Health protection | 44 |
| Health improvement | 97 |
| Mental health | 14 |
| Integrated care | 10 |
| Wider determinants of health | 54 |
| Self-management support | 26 |
| Global health | 10 |
| Epidemiology | 23 |
| Early diagnosis, detection and intervention | 17 |
| Prevention in health policy, strategies and care pathways | 20 |
| Secondary prevention | 20 |
| Public health models and services | 69 |

Educators’ open-text responses add context to the above data as often the summaries of data may inadvertently make assumptions about practice. The quantitative data showed that educators want more guidance, yet the use of frameworks was very low. Respondents subsequently highlighted that the utility of guidance is key:

> *“Most of the frameworks are long documents consisting of many pages and there is limited time to complete reading about these, so provision of a summary would be very much helpful*.*” (R110)*

Likewise educators’ descriptions of what content is covered cannot be equated with absence of all relevant teaching. The reporting of topics and formats does not demonstrate how every learner may encounters them:

> *“I think we do deliver a wide range of preventative health within the curricula but may not clearly identify it as such” (R89)*

The format of current education showed that weaving content through curricula was a common approach across the shifts (Figure 3).

**Figure 3.**
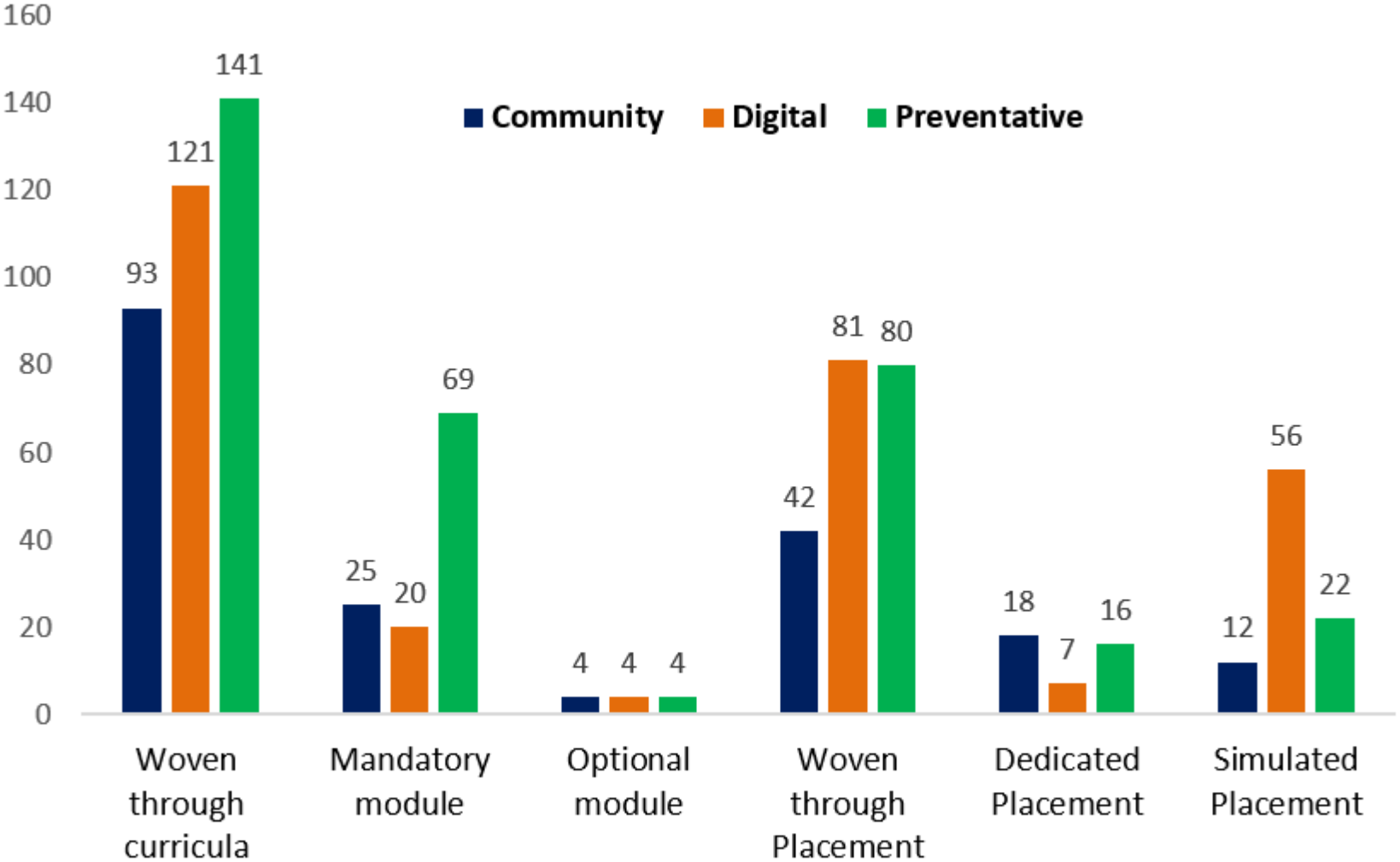
Current teaching format of each shift.

### Integrated interpretation of findings

Table 3 brings together quantitative findings, open-text categories, and interpretations. Digital provision raised questions about educators’ expertise and access to technology; community provision about placements and service partnerships; and prevention about the breadth and depth of learning within crowded curricula. These are accounts of experienced or anticipated challenges and existing or possible support, rather than demonstrated causes of the priority ratings.

**Table 3.** Joint display of survey findings.

| **Domain** | **Finding** | **Integrated interpretation** |
|---|---|---|
| Digital provision | Growing importance not yet reflected | Practical support for educators and access to technology may help develop teaching. |
| Community provision | Included with plans to do more | Placement opportunities are important for curriculum development |
| Prevention provision | Mature but not consistent | Prevention was often included, but support still needed |
| Use of guidance | Very limited use of frameworks or national support | Resource availability does not predict awareness, use, or impact |

## Discussion

The current research describes educators’ accounts of pre-registration education relevant to England’s NHS three shifts. It also identifies challenges experienced or anticipated, and support that helped or could help develop and sustain future fit provisions.

Overall the maturity of education relating to the three shifts education was low with fewer than a third of respondents describing prevention as a strong component, despite it being the most developed area. Considered alongside the open text data, the findings suggest that educators need specialist expertise, clearer guidance from regulatory bodies, and local support.

The digital findings are relevant beyond England. In Australia, it has also been highlighted that educators are crucial to equipping future practitioners to respond to the rapidly changing digital health landscape. The authors developed a nursing artificial intelligence curriculum framework noting the current gaps in education content, educator workforce expertise, and preparedness of graduates [23], verifying the findings of the current research.

Community education depends partly on the learning opportunities that local systems can offer. Placement difficulties and requests for collaboration in the current research suggest that this area is fundamental to neighbourhood models which can support learning across professional and organisational boundaries [7]. This dominance of hospital-based placement opportunities, and emerging need for non-traditional placements, is echoed across international literature. Despite the clear mandate by the WHO for educational institutions to direct education to meet the needs of their locality, place-based and community placements continue to be secondary to hospital experiences, even in programmes designed to target local workforce development and embedded training [24].

For prevention, contemporary professional standards and public health initiatives offer a foundation [12,13], but educators in the current research still raised concerns about breadth, curriculum space, workforce knowledge and content relevance. The Council of Deans’ earlier work similarly found that prevention education is taking place while practical support is still needed [12]. Others have written about the fallacy in healthcare education where skills, tasks, a biomedical heavy paradigm and inability to develop consensus around the art and science of professions hinder the inclusion of elements like poetry, digital health, and primary prevention [25, 26].

Low reported framework and national resource use should prompt an investigation into the usefulness and implementation of resources, rather than the development of more frameworks. Requests for shorter guidance and clearer expectations suggest a practical role for regulators and professional bodies to help programmes distinguish what students need for entry to practice from learning. Rather than prescribing uniform checklists, co-developed examples and explicit links between learning outcomes and practice may provide a more useful basis for curriculum development across professions. Other authors have argued that a continued attempt to list competencies fails to recognise that professions are historical tribes that will not inherently change due to increasing volumes of tasks and that a more nuanced ethically and values-led set of principles for changing healthcare should be given primary [27, 28].

Interprofessional education offers one approach to exploring the intersections between the shifts [29,30]. The request for interprofessional nontraditional placements offers a concrete example of the opportunities educators would like to develop. Learning involving community partners could combine prevention, coordination and digital care in the same context. Drawing on these findings, future studies should examine both implementation and transfer into practice, including observation or assessment of performance and perspectives from learners, supervisors and service users.

Longitudinal follow-up through pre-registration to early employment would help distinguish educational gains from the support and constraints encountered through education.

These findings also raise questions about what students have the opportunity to develop beyond familiarity with curriculum content. Accounts of limited time to develop skills and the need to focus of portable job-ready elements suggest a useful direction for further enquiry. Capability may offer one way to explore how students learn to apply their knowledge in changing or unfamiliar situations [31, 32].

### Strengths and limitations

The findings offer key learning about implementation beyond England, particularly where institutions face similar gaps between national priorities and local educational resources. The study included educators from all key professions (except art therapy) across all English regions and considered the three shifts within one questionnaire. Bringing open-text responses alongside numerical patterns enabled authors to move beyond identifying whether three shifts-related content was present, providing insight into what educators described as already aligning with the three shifts, as well as the challenges to further development and potential ways of addressing them. However, recruitment through networks was voluntary, and the proportion of uptake versus invitation is unknown. Respondents also differed in their responsibilities, how much of the curriculum they could describe, and had small counts from some AHPs. The sample should therefore not be considered nationally representative.

Reports rely on accounts from educators rather than curriculum audits, observations of teaching or student outcomes. The priority categories combine importance and current provision, and the lists of named resources differ in length and relevance between shifts and professions. Short written responses provide limited detail, and people who contributed text may differ from those who answered only fixed-response questions. Our comparison of grouped findings cannot link an individual’s rating to their explanation or establish cause and effect. These limitations restrict what we can conclude about readiness or the effectiveness of educational approaches.

## Conclusion

Educators reported uneven provision relevant to the NHS three shifts, with digital health least often described as a strong component. The open-text accounts suggest that further development requires attention to curriculum space, educator expertise, technologies and practice-learning opportunities. The distinction between introducing “concepts and ideas” and developing skill, illustrated by the prevention findings, captures an important educational concern: content inclusion alone does not establish what students can do. Universities, services and regulators have complementary roles in making meaningful learning possible. Further assessment and follow-up are needed to establish whether educational changes prepare students for future practice.

## Data Availability

Data is available upon reasonable request from the lead author

## Declarations

## Data availability

Anonymised data can be requested from the lead author via the following.

## Acknowledgements and contributions

The team would like to thank the participants, our expert reference group, and colleagues from NHS England, professional bodies and healthcare regulators.

## Related outputs

This survey is part of a wider programme of work that will be published in the near future as a grey literature report and subsequent peer reviewed papers.

## Writing assistance

Artificial intelligence was not used in the preparation of this manuscript.

